# Analysis of all video-assisted thoracic sympathectomy over 9 years in Brazil: trends, costs and mortality over 200 million inhabitants

**DOI:** 10.1101/2025.11.24.25340923

**Authors:** Bruno Jeronimo Ponte, Jose Renato Saback Fonseca, Alexandre Fioranelli, Marcelo Fiorelli Alexandrino da Silva, Marcelo Passos Teivelis, Jose Ribas Milanez de Campos, Edson Amaro, Nelson Wolosker

## Abstract

**Background:** Hyperhidrosis (HH) is characterized by excessive sweating beyond thermoregulatory requirements and can significantly impair quality of life. Although oxybutynin has become an effective first-line therapy, video-assisted thoracic sympathectomy (VATS) remains the only definitive option for refractory cases. The availability of nationwide public and private healthcare datasets in Brazil enables a comprehensive assessment of national procedural trends on a continental scale.

**Methods:** We performed a retrospective nationwide analysis of all VATS procedures for HH conducted in Brazil between 2015 and 2023. Data were extracted from publicly available governmental databases covering both the public and private healthcare systems. Procedure rates were evaluated by year, geographic region, sex, and age group. Temporal patterns and differences between healthcare sectors were analyzed.

**Results:** A total of 26,980 VATS procedures were performed during the study period, with the private sector accounting for 75.25% of surgeries. Regionally, the Southeast (48.28%) and South (29.51%)—the most economically developed areas of the country—concentrated the majority of procedures. Women represented 64.5% of operated patients, and most surgeries were performed in individuals aged 15–39 years (82.74%). The mean cost per procedure in the private system was 4.2 times higher than in the public system (US$1,351.94 vs. US$351.27, p < 0.001), and the private sector accounted for more than 90% of total national expenditures. The overall mortality rate was 0.048%, with no significant difference between healthcare sectors.

**Conclusions:** This study provides the first nationwide evaluation of VATS performed for HH across Brazil’s public and private systems. Based on 26,980 procedures carried out between 2015 and 2023, most surgeries occurred in the private sector and predominantly involved women and young adults aged 15–39 years. Although mortality was low and similar across systems, procedure costs were substantially higher in the private sector, which accounted for the vast majority of national expenditures.

## Introduction

Hyperhidrosis (HH) is a condition marked by excessive sweating that exceeds the body’s physiological requirements for temperature regulation. (1,2) The global prevalence of HH varies considerably, with estimates ranging from 2.8% to 12.76%. (3–7). Symptoms usually begin during childhood and are more common in women.(8) The sites most frequently affected include the palms, axillae, soles of the feet, and face, often impacting multiple regions simultaneously. (2) The pathophysiology is not fully understood but is thought to result from excessive stimulation of the sympathetic nervous system at its regulatory center. Conditions or emotional factors with a psychosomatic component often trigger or worsen HH, enhancing autonomic activity and increasing sweat production. (9,10)

Until the end of the last decade, the initial treatment for HH patients was video assisted thoracic sympathectomy (VATS). (9,11,12) However, recent studies have demonstrated the effectiveness and safety of oxybutynin, an anticholinergic medication, which provides clinical improvement in over 70% of users.(6,7) Since this discovery, we have adopted oxybutynin as the first-line treatment option.(13–15) For patients who do not respond adequately to this medication, VATS, the only definitive treatment, is now considered the gold standard for the definitive treatment of HH.(11,16,17) This procedure significantly enhances patients’ quality of life and shows good results concerning several factors, including body mass index, resection level, preoperative quality of life, and the number of resected ganglia.(8,18–20)

Other effective alternatives for treating HH include the use of glycopyrronium, (21) and botulinum toxin injections. (22) However, one point remains clear: without appropriate treatment, patients may experience significant psychosocial and occupational impairment, leading to a marked reduction in quality of life. (23,24)

Brazil has a population of approximately 200 million people (25) 72% of the population depends on the publicly funded Unified Health System (SUS), while most of the remaining citizens are covered by employer-sponsored private health insurance plans (about 23%). A small minority (less than 5%) pays out of pocket for health services. (26)

Brazil’s public healthcare system has developed a digital platform that collects data on procedures performed in public hospitals nationwide. More recently, the private healthcare sector has also begun making its data available to the public, adopting practices like those of the public sector. The integration of both datasets now allows for a comprehensive national assessment of healthcare, something previously hindered by the country’s vast territory and limited data availability.

To the best of our knowledge, this is the first study to describe and analyze such a large-scale dataset in a country of continental dimensions. The results offer a vision of the total, and also comparative evidence that can guide the development of management strategies for HH specifically adapted to the Brazilian context. (27)

This study aimed to analyze nationwide data from both the public and private healthcare systems in Brazil between 2015 and 2023 regarding VATS for HH. The objectives were to evaluate VATS rates and to identify differences across regions, age groups, and sexes within both the public and private healthcare sectors.

## Methods

This retrospective, cross-sectional, population-based study included all patients diagnosed with HH who underwent VATS in Brazil between 2015 and 2023. Data from both public and private healthcare systems were analyzed to provide a comprehensive national overview. The study was exempt from ethics committee approval because it used publicly available data and was registered under protocol 10337 by the institution’s research department. All data are anonymized and accessible through government platforms; therefore, informed consent was waived.

Patients were divided into two groups: those from the public healthcare system and those from the private healthcare system. The public included all patients treated through the SUS, while the private included those with health insurance coverage.

Data were extracted from the TabNet platform of the Department of Informatics of the Unified Health System (DATASUS) and from the Private Sector Database (D-TISS). (28–30) DATASUS compiles information on procedures performed in public hospitals across Brazil, where notification is required for the procedure to be eligible for payment. The D-TISS is managed by a private system that grants access to procedure data. Overall, population samples were derived from the Brazilian Institute of Geography and Statistics (25), as well as DATASUS and D-TISS. (28–30) The STROBE guidelines were duly followed and respected. (31)

This study analyzed VATS procedures performed in Brazil over 9 years, encompassing both public and private health systems and covering almost all national services. The extracted data included the prevalence of sex (biological assignment at birth), patient age, number of procedures performed in-hospital, and geographic region. The number of VATS per region was extracted and stratified by the region’s population. VATS procedures were identified in DATASUS by code 0403050146 and in D-TISS code 31403379.

We initially estimated the total number of sympathectomies performed in public and private healthcare services from 2015 to 2023. Next, we assessed the distribution of these procedures by gender and age group within the Unified Health System (SUS) and Private Healthcare Services (PHS). We also analyzed the regional distribution of procedures across Brazil, as well as the procedure rate per one million individuals. Following this, we examined the investments made in SUS and PHS during that period, including the average cost per procedure by year and funding source. Finally, we evaluated the total number of deaths among patients undergoing VATS in Brazil.

Statistical analyses were performed using SPSS software, version 20.0 (IBM Corp., Armonk, NY). Associations between procedure rates, reimbursement values, year, healthcare sector, and region were evaluated using generalized linear models with a Gamma distribution. Models with a Poisson distribution were applied to analyze age- and sex-specific rates, incorporating the number of procedures as an offset. Comparisons of mortality between sectors were performed using Fisher’s exact test due to the low number of deaths. Results were expressed as rate or mean ratios with corresponding 95% confidence intervals, and a p-value < 0.001 was considered statistically significant.

## Results

Table 1 shows the number and proportion of VATS procedures performed in Brazil under the public and private systems between 2015 and 2023. It also presents rates per million. A total of 26,980 VATS procedures were performed, of which 6,678 (24.75%) were performed in the public and 20,302 (75.25%) in the private. Over the years, the majority of procedures were done through the private system (66 to 84%). The absolute number of surgeries fell in 2020 and 2021. There was a decline in the public system from 33.97% in 2015 to 20% in 2023.

**Table 1.**
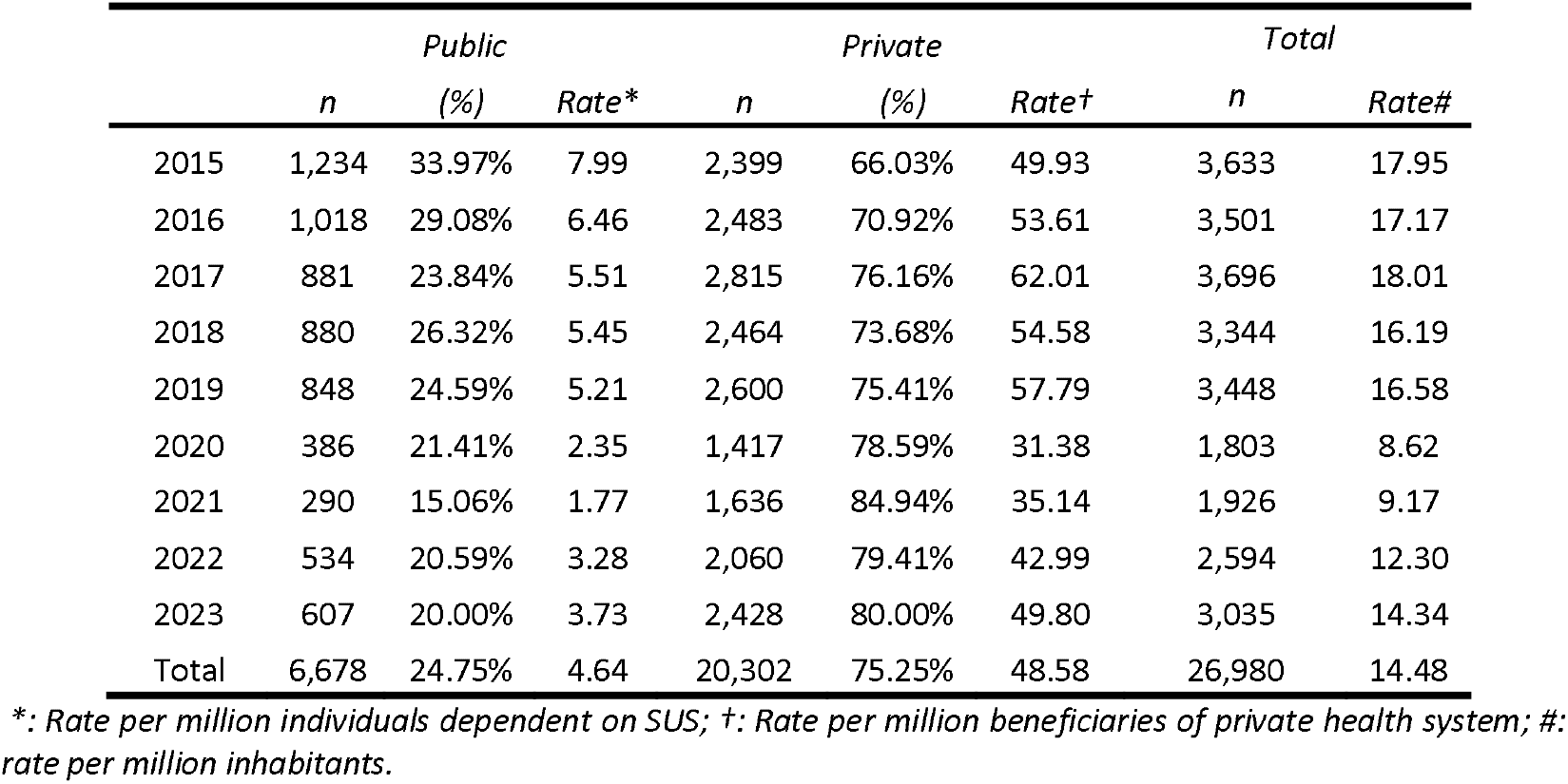
Number of video-assisted thoracoscopic sympathectomies performed in the SUS and PHS from 2015 to 2023.

Table 2 presents the average ratios (per 1,000,000 individuals) with 95% confidence intervals and p-values. The adjusted model shows a yearly decline in VATS rates of 6.3% (0.937; 95% CI 0.903–0.973; p = 0.001). Compared to the public system, the private displays a higher rate (12.299; CI 10.038–15.070, p < 0.001). Considering Southeast as reference, the Midwest (0.822; 95% CI 0.600–1.125, p = 0.220), Northeast (0.663; 95% CI 0.482–0.911, p = 0.011), and North (0.178; 95% CI 0.130–0.243, p < 0.001), show lower rates, while the South (2.084; 95%CI 1.526 – 2.847 p<0.001) shows a higher one.

**Table 2.** Procedure rate for 1,000,000 individuals.

|  | **Average Ratio (CI 95%)** | ***p value*** |
| --- | --- | --- |
| Year | 0.937 (0.903; 0.973) | 0.001 |
| Source |  |  |
| Public | Reference |  |
| Private | 12.299 (10.038; 15.070) | <0.001 |
| Region |  |  |
| Southeast | Reference |  |
| Midwest | 0.822 (0.600; 1.125) | 0.220 |
| Northeast | 0.663 (0.482; 0.911) | 0.011 |
| North | 0.178 (0.130; 0.243) | <0.001 |
| South | 2.084 (1.526; 2.847) | <0.001 |

*CI: Confidence interval*

In the public system, 64.5% of procedures were performed on women (4,307). In the private network, 56.7% were women (11,502), 41.6% were men (8,450), and 1.7% were individuals whose sex was not reported. Considering only cases with reported sex, women comprised 57.6%. A statistically significant difference in sex distribution was observed between the two healthcare systems (p < 0.001), demonstrating a higher prevalence of procedures among female patients.

Table 3 shows the distribution of procedures by age in public and private systems. Younger age groups (<14 and 15–19 years) were more frequent in the public system. In older groups (20–49 years), the private system had a higher proportion, with statistical significance (p < 0.001).

**Table 3.** Distribution of procedures by age groups between public and private healthcare systems.

| Age Group<br>(years-old) | Public |  | Private |  | Total |  |
| --- | --- | --- | --- | --- | --- | --- |
|  | *n* | (%) | *n* | (%) | *n* | (%) |
| **<14** | 592 | 8.86% | 1,275 | 6.28% | 1,867 | 6.92% |
| **15-19** | 2,020 | 30.25% | 3,889 | 19.16% | 5,909 | 21.90% |
| **20-29** | 2,359 | 35.32% | 7,832 | 38.58% | 10,191 | 37.77% |
| **30-39** | 1,136 | 17.01% | 5,088 | 25.06% | 6,224 | 23.07% |
| **40-49** | 356 | 5.33% | 1,444 | 7.11% | 1,800 | 6.67% |
| **50-59** | 118 | 1.77% | 301 | 1.48% | 419 | 1.55% |
| **60-69** | 70 | 1.05% | 89 | 0.44% | 159 | 0.59% |
| **70-79** | 21 | 0.31% | 29 | 0.14% | 50 | 0.19% |
| **>80** | 6 | 0.09% | 5 | 0.02% | 11 | 0.04% |
| **Not informed** | 0 | 0.00% | 350 | 1.72% | 350 | 1.30% |
| **Total** | 6,678 | 100% | 20,302 | 100% | 26,980 | 100% |

Table 4 shows the distribution of procedures across Brazilian regions within the public and private systems. In all regions, the private system accounts for most procedures (68.5% to 77.6%). The Southeast region has the largest share of total procedures (48.28%). Regions with lower population density, such as the Midwest (7.29%) and North (0.89%), reported the lowest procedure rates.

**Table 4.** Procedures distribution between Brazilian regions.

| Region | Public |  |  | Private |  |  | Total |  |  |
| --- | --- | --- | --- | --- | --- | --- | --- | --- | --- |
|  | *n* | (%) | Rate* | *n* | (%) | Rate† | *n* | (%) | Rate# |
| **North** | 76 | 31.54% | 5.96 | 165 | 68.46% | 87.15 | 241 | 0.89% | 16,47 |
| **Northeast** | 847 | 22.38% | 17.08 | 2,938 | 77.62% | 405.81 | 3,785 | 14.03% | 66,62 |
| **Midwest** | 485 | 24.66% | 29.99 | 1,482 | 75.34% | 412.70 | 1,967 | 7.29% | 99,54 |
| **Southeast** | 2,928 | 22.48% | 49.62 | 10,098 | 77.52% | 331.03 | 13,026 | 48.28% | 145,52 |
| **South** | 2,342 | 29.42% | 99.44 | 5,619 | 70.58% | 765.55 | 7,961 | 29.51% | 257,72 |

\*: Rate per million individuals dependent on SUS; †: Rate per million beneficiaries of private health system; #: rate per million inhabitants

Table 5 presents the total financial investments for each healthcare system. From 2015 to 2023, combined expenditures from both the public and private sectors totalled U$29,499,445.19. The public allocated U$2,321,056.69 for VATS, while the private invested U$ 27,178,388.50.

**Table 5.** Investments made in SUS and PHS between 2015 and 2023.

| Year | Public | | | Private | | | Total (U\$) |
| --- | --- | --- | --- | --- | --- | --- | --- |
| | n (U\$) | % | Per Procedure (U\$) | n (U\$) | % | Per Procedure (U\$) | |
| 2015 | 436,917.56 | 15.46% | 354.07 | 389,778.26 | 84.54% | 996.16 | 2,826,695.82 |
| 2016 | 348,401.77 | 10.62% | 342.24 | 2,931,646.24 | 89.38% | 1,180.69 | 3,280,048.01 |
| 2017 | 306,075.48 | 7.92% | 347.42 | 3,559,972.34 | 92.08% | 1,264.64 | 3,866,047.82 |
| 2018 | 292,205.77 | 8.52% | 332.05 | 3,137,841.89 | 91.48% | 1,273.47 | 3,430,047.66 |
| 2019 | 283,138.52 | 7.40% | 333.89 | 3,542,394.56 | 92.60% | 1,362.46 | 3,825,533.07 |
| 2020 | 147,797.51 | 6.98% | 382.90 | 1,971,143.36 | 93.02% | 1,391.07 | 2,118,940.88 |
| 2021 | 106,812.30 | 4.19% | 368.32 | 2,441,277.10 | 95.81% | 1,492.22 | 2,548,089.40 |
| 2022 | 186,574.69 | 5.42% | 349.39 | 3,255,529.06 | 94.58% | 1,580.35 | 3,442,103.75 |
| 2023 | 213,133.10 | 5.12% | 351.13 | 3,948,805.68 | 94.88% | 1,626.36 | 4,161,938.78 |
| Total | 2,321,056.69 | 7.87% | 351.27 | 27,178,388.50 | 92.13% | 1,351.94 | 29,499,445.19 |

Table 6 presents the average value per procedure by year and source. The adjusted model for average VATS cost shows a yearly increase of 3.1%. In addition, average reimbursement in the private sector is estimated at 4.2 times that of the public sector.

**Table 6.** Average value per procedure by year and source.

| □ | Average ratio (CI 95%) | p value |
| --- | --- | --- |
| Year | 1.031 (1.016; 1.045) | <0.001 |
| Source |  |  |
| Public | Reference |  |
| Private | 4.203 (3.915; 4.513) | <0.001 |

Table 7 shows the absolute distribution of deaths among patients who underwent VATS in Brazil between 2015 and 2023. A total of 10 deaths were recorded, representing an overall mortality rate of 0.048%. Mortality associated with VATS remained very low, with five deaths among 6,678 VATS procedures in public and five among 20,302 in the private, corresponding to 749 and 246 deaths per million procedures, respectively. There was no statistical difference between systems, although the odds ratio for death in public versus private settings was 3.04 (95% CI, 0.70 to 13.21).

**Table 7.** Absolute distribution of deaths of patients undergoing VATS in Brazil.

| Year | Public | Private | Total |
| --- | --- | --- | --- |
| 2015 | 1 | 0 | 1 |
| 2016 | 1 | 0 | 1 |
| 2017 | 0 | 3 | 3 |
| 2018 | 1 | 1 | 2 |
| 2020 | 2 | 0 | 2 |
| 2023 | 0 | 1 | 1 |
| Total | 5 | 5 | 10 |

## Discussion

This retrospective, cross-sectional study encompassed most of the Brazilian population, analyzing a comprehensive sample of 26,980 VATS performed nationwide between 2015 and 2023. The analysis included both public and private healthcare systems, providing a broad and representative overview of surgical practice across diverse socioeconomic and geographic contexts. To date, this study represents the longest and first nationwide cohort evaluating VATS for HH, offering robust epidemiological insight into the distribution, accessibility, and outcomes of definitive surgical treatment for HH throughout Brazil.

Data were obtained from national databases—DATASUS, which records all publicly funded procedures, and D-TISS, which compiles private healthcare data—ensuring near-complete national coverage (excluding only out-of-pocket cases), as reporting is mandatory for reimbursement. Additionally, population data from the Brazilian Institute of Geography and Statistics (IBGE) and complementary epidemiological sources were integrated, allowing precise demographic stratification and the calculation of standardized rates that strengthen the validity and comparability of the findings.

Over nine years, a total of 26,980 VATS were performed in Brazil, mainly in the private system (75.25%). Annual volumes in the private setting historically hovered around 2,500 cases, while the public sector contributed approximately 900 procedures. However, in 2020– 2021, these figures declined to nearly 1,500 in the private sector and just 300 in public hospitals, a downturn likely associated with the COVID-19 pandemic. (32) Since then, the numbers have partially rebounded, but not yet to pre-pandemic levels—perhaps in part due to evolving HH treatment strategies in Brazil. Today, many patients seeking care for HH are first managed medically with oxybutynin, which leads to clinical improvement for a substantial fraction and may reduce the necessity for surgical intervention.(1,21)

There is no comprehensive nationwide database to reliably enumerate VATS for HH. Available reports stem from institutional or regional case series (e.g., University of Maryland, Barrow Neurological Institute) and lack full national coverage. In contrast, Brazil remains unique in having a national epidemiological account of all VATS, derived from its public and private health system databases. In other countries, such as Taiwan or in European nations, data are usually limited to high-volume centers or regional registries, again lacking nationwide scope. (4,33) Because of this, no study to date has comprehensively used U.S. administrative hospital databases (e.g., NIS or SID) to estimate the total burden of thoracic sympathectomy for HH across the country.

Although such databases are often used in epidemiologic research, they encounter limitations in distinguishing surgical indications, capturing outpatient or outpatient-like procedures, and reliably coding HH as the operative rationale. Therefore, future methodological work is needed to develop and validate algorithms (based on diagnosis and procedure codes) to identify sympathectomy for HH in these large databases—and thereby enable cross-national comparisons of surgical practice over time.

Our data reveal significant differences between the public and private sectors. In the public, the rates ranged from 15.06% to 33.97%. After adjusting for population size, VATS rates remained higher in the private sector, though the gap has increased, and the private system now performs VATS at an estimated rate 12 times that of the public system. (17,34)

This disparity suggests that patients in the public system have longer wait times than those in the private system, and this may also be attributed to factors such as personal preferences or barriers to accessing surgery. (34)

Consistent with previous reports, there was a predominance of female patients undergoing VATS. (17) While the prevalence of HH is similar between sexes, women tend to seek treatment more frequently, partly due to aesthetic and psychosocial concerns. (35) In a public-only Brazilian series, women represented about 65% of patients undergoing sympathectomy. (17,35) Similarly, a multicenter clinical trial reported that 63.5% of patients undergoing surgery for HH were female. (12) Added to that, in our present study, 57.6% of patients were female, with a statistically significant difference observed in both sectors (p < 0.001).

In terms of age, HH is recognized as a condition that affects younger individuals, often manifesting during childhood or adolescence, yet many patients delay treatment until young adulthood. (18) In a multicenter clinical trial, the mean age of presentation was 24 years. (12) In Brazilian data from São Paulo, 66.2% of VATS were performed on patients aged 20-39 years, (35) and in national SUS analyses, more than 80% of patients fell between 15 and 59 years old. (36) In Taiwan, the highest prevalence rates were observed in the 20-29 age group. (33)

Those with HH generally seek care to improve their quality of life;(13) however, many wait until symptoms intensify, often during their economically active years or occasionally when prompted by family during childhood. This results in prolonged periods of personal, social, and psychological burden before access to treatment is achieved. (9,16)

In our dataset, we observed that younger patients (15–29 years) were predominantly treated within the public sector, whereas the private sector exhibited a greater proportion of older patients (20-39 years). This pattern may reflect differential access to early, non-surgical therapies (e.g. topical, oral, or minimally invasive treatments) in the private system, delaying or reducing the need for surgery in younger individuals. Conversely, adults in private care may have greater awareness, financial resources, and access to specialized centers, leading them to opt for surgical intervention more readily. These dynamics suggest a complex interplay between disease natural history, patient preferences, and structural access to care.

Regional analysis revealed that VATS procedures in the private sector significantly exceeded those performed in the public sector, particularly in the Southeast and South regions, where 77.97% of all procedures were concentrated. This should be explained by the fact that these regions possess the country’s highest economic productivity, population density, and Human Development Index (HDI), as well as a more robust private healthcare sector compared with other parts of Brazil. Overall, VATS rates per one million inhabitants were markedly higher in the South and Southeast, while lower rates were observed in the North, a pattern that likely reflects underlying disparities in health policy, hospital infrastructure, and patient demographics.(37,38)

Another key contributing factor is the uneven distribution of thoracic surgeons, which further accentuates regional variation in procedure rates. The South had the highest density of thoracic surgeons, with 9.14 per million inhabitants, followed by the Southeast (7.10), the Midwest (5.32), the Northeast (3.65), and the North (2.80). Thus, regions with higher economic capacity, HDI, and surgeon density were responsible for the largest VATS volumes. (39)

When analyzing the funds allocated and reimbursed for VATS in private and public systems between 2015 and 2023, we observed that the private sector accounted for over 92% of total investment in these years, highlighting a profound imbalance in financial engagement. Moreover, the per-procedure reimbursement in the private sector is approximately 4.2 times higher than in the public system, according to the adjusted cost model.

This disproportion suggests that public institutions may be structurally undercompensated for these procedures, risking underfunding or cost absorption by hospitals. The estimated yearly increase of 3.1 % in average VATS cost further compounds this gap, as inflation, technological advances, and rising input costs progressively erode public sector margins. (37) The dominance of private investments also points to potential inequities in access; patients with private coverage or financial means are more likely to receive interventions, while those dependent on public may face delayed or restricted access to care. (38) Overall, these financial dynamics underscore how structural funding and reimbursement policies can directly influence procedural volume, care quality, and equitable distribution of advanced vascular or thoracic services in Brazil.

The mortality related to VATS procedures in Brazil between 2015 and 2023 exhibited an exceptionally low death rate of 10 fatalities across all cases, yielding an overall mortality of just 0.048%. There were five deaths in public over 6,678 procedures and five deaths in private over 20,302 procedures, suggesting a numerically higher mortality rate in the public system (749 deaths per million) compared to the private (246 deaths per million). However, this difference did not reach statistical significance. The estimated odds ratio (3.04; 95 % CI 0.70–13.21) hints at a trend toward greater relative risk in the public system, but the wide confidence interval underlines the uncertainty arising from the small number of events.

These results align with the perception that VATS is a safe minimally invasive modality when performed at experienced centers, reflected in international data showing low surgical mortality for thoracoscopic procedures. The same was observed in previous studies. (17) In a 2004 study, nine perioperative deaths were described: five from massive intrathoracic hemorrhage, three from anesthetic complications during mechanical ventilation, and one of undetermined cause. (40) Other retrospective and prospective cohorts reported no mortality following. (9,16)

Mortality is a crucial parameter to assess in any form of medical treatment; however, on its own, it does not capture the full range of surgical risk — including major complications, conversion to open surgery, length of hospital stays, and long-term outcomes — which were not studied in this work because such data were unavailable in the databases.

## Limitations

The study’s cross-sectional design precludes causal inferences, limiting interpretations to associations and temporal patterns. The absence of patient-level clinical data — such as comorbidities, disease stage, quality of life, and reasons for choosing VATS—further constrains the analysis of specific risk factors and clinical contexts. Use of aggregated administrative databases (DATASUS and D-TISS) also introduces inherent limitations, including potential coding errors, reporting inconsistencies, and underreporting. Because both sources are anonymized, longitudinal follow-up and assessment of outcomes or comorbidities were not feasible.

Despite these limitations, this is the first nationwide analysis of VATS procedures in both public and private healthcare systems, offering real-world data with high external validity and highlighting significant differences in treatment volume and financial investment between the two sectors.

## Conclusions

Based on all 26,980 VATS performed in Brazil between 2015 and 2023 — corresponding to an annual rate of 14.48 per million inhabitants — we conclude that most procedures were carried out in the private system (75.25%, 12.3 times more than in the public sector). Most procedures were performed on women (64.5%) and predominantly on young individuals aged 15–39. The mean cost per procedure in the private system was 4.2 times higher than in the public system (U$1,351.94 vs. U$351.27), with the private sector being responsible for over 90% of total expenditures. The overall mortality rate was very low (0.048%), with no significant difference between the two systems.

## Data Availability

All data produced in the present study are available upon reasonable request to the authors

https://datasus.saude.gov.br/informacoes-de-saude-tabnet

